# How different pre-existing mental disorders and their co-occurrence affects clinical outcomes of COVID-19? A study based on real-world data in the Southern United States

**DOI:** 10.1101/2021.10.21.21265340

**Authors:** Shan Qiao, Jiajia Zhang, Shujie Chen, Bankole Olatosi, Suzanne Hardeman, Meera Narasimhan, Larisa Bruner, Abdoulaye Diedhiou, Cheryl Scott, Ali Mansaray, Sharon Weissman, Xiaoming Li

## Abstract

**Importance:** A growing body of research focuses on the impact of pre-existing mental disorders on clinical outcomes of COVID-19 illness. Although a psychiatric history might be an independent risk factor for COVID-19 infection and mortality, no studies have systematically investigated how different clusters of pre-existing mental disorders may affect COVID-19 clinical outcomes or showed how the coexistence of mental disorder clusters is related to COVID-19 clinical outcomes.

**Objective:** To explore how different pre-existing mental disorders and their co-occurrence affects COVID-19-related clinical outcomes based on real-world data.

**Design, Setting, and Participants:** Using a retrospective cohort study design, a total of 476,775 adult patients with lab-confirmed and probable COVID-19 between March 06, 2020 and April 14, 2021 in South Carolina, United States were included in the current study. The electronic health record data of COVID-19 patients were linked to all payer-based claims data through the SC Revenue and Fiscal Affairs Office.

**Main Outcomes and Measures:** Key COVID-19 clinical outcomes included severity, hospitalization, and death. COVID-19 severity was defined as asymptomatic, mild, and moderate/severe. Pre-existing mental disorder diagnoses from Jan 2, 2019 to Jan 14, 2021 were extracted from the patients’ healthcare utilization data via ICD-10 codes. Mental disorders were categorized into internalizing disorders, externalizing disorders, and thought disorders.

**Results:** Of the 476,775 COVID-19 patients, 55,300 had pre-existing mental disorders. There is an elevated risk of COVID-19-related hospitalization and death among participants with pre-existing mental disorders adjusting for key socio-demographic covariates (i.e., age, gender, race, ethnicity, residence, smoking). Co-occurrence of any two clusters was positively associated with COVID-19-related hospitalization and death. The odds ratio of being hospitalized was 2.50 (95%CI 2.284, 2.728) for patients with internalizing and externalizing disorders, 3.34 (95%CI 2.637, 4.228) for internalizing and thought disorders, 3.29 (95%CI 2.288, 4.733) for externalizing and thought disorders, and 3.35 (95%CI 2.604, 4.310) for three clusters of mental disorders.

**Conclusions and Relevance:** Pre-existing internalizing disorders, externalizing disorders, and thought disorders are positively related to COVID-19 hospitalization and death. Co-occurrence of any two clusters of mental disorders have elevated risk of COVID-19-related hospitalization and death compared to those with a single cluster.

## INTRODUCTION

Outbreaking in late 2019, the coronavirus disease 2019 (COVID-19) caused by the severe acute respiratory syndrome coronavirus (SARS-CoV-2) has rapidly become a global public health crisis. The World Health Organization (WHO) declared COVID-19 a pandemic on March 11, 2020 (1). The COVID-19 pandemic has continued to evolve causing a tremendous death toll and severe somatic complications in millions globally. People who have existing mental disorders including internalizing disorders (e.g., depression, generalized anxiety disorder, panic disorder, and posttraumatic stress disorder), externalizing disorders (e.g., conduct disorder, alcohol dependence, cannabis dependence, other drug dependence, and tobacco addiction), and thought disorders (e.g., obsessive-compulsive disorder, mania, and schizophrenia) may be particularly vulnerable(2-8). They may be more susceptible to contracting COVID-19 and show general deterioration of mental health (2-8). Studies show patients with suspected and diagnosed psychiatric issues have shown elevated psychiatric distress, additional anxiety symptoms, poor sleep, and quality of life concerns during the COVID-19 pandemic (9, 10). In addition, the COVID-19 pandemic may impede the physical health of people with mental disorders because of reduced or interrupted access to healthcare services. This change in access may undermine the management of chronic diseases and exacerbate psychiatric disorders (11, 12). Also, physical distancing requirements during the pandemic may lead to decreased social support and further worsen mental health status (13).

A growing body of research focuses on the impact of pre-existing mental disorders on clinical outcomes of COVID-19 illness among this vulnerable population. According to cohort studies in South Korea and United Kingdom (UK), pre-existing mental disorders were associated with risk of severe COVID-19 clinical outcomes (hospitalization, intensive care unit [ICU] admission, invasive ventilation, or death) after adjusting for age, sex, ethnicity, somatic comorbidities, and regional COVID-19 influential factors (14-17). Recently, a medical record review study and a large electronic health record (EHR) case-control study in the United States found a higher mortality rate among COVID-19 patients with mental disorders compared to the general population (18, 19). Similarly, an Italian cross-sectional study among patients hospitalized with COVID-19 showed patients with severe psychiatric disorders died at a younger age than those without a psychiatric disorder after controlling for other clinically relevant variables.

Although existing literature suggests that a psychiatric history might be an independent risk factor for COVID-19 infection and mortality, there are still some knowledge gaps worth further exploration. First, no studies have systematically investigated how different clusters of pre-existing mental disorders may affect COVID-19 clinical outcomes (20). Increasing evidence reveals that sets of mental disorders and symptoms predictably co-occur (21, 22). That is, some mental disorders are more highly correlated with each other. Studies about the structure of psychopathology result in the incorporation of disorder clusters in research and the growth of transdiagnostic treatment (23, 24). For example, according to an existing psychopathology structure model, i.e., “three factor model”(25), mental disorders can be organized into three broad clusters (or higher-order factors): “internalizing disorders” (people with internalizing disorders keep maladaptive emotions and cognitions to themselves or internalize problems, e.g., depression and anxiety), “externalizing disorders” (mental disorders characterized by externalizing behaviors, maladaptive behaviors directed toward an individual’s environment, which cause impairment or interference in life functioning, e.g., antisocial and substance use disorders), and “thought disorders” (disturbance in cognition that adversely affects language and thought content, and thereby communication, e.g., schizophrenia and schizotypal personality disorders) (25-27). Examining the different roles of these mental disorder clusters in affecting COVID-19 clinical outcomes will advance our understanding of the intersection of mental disorders and infectious diseases in the context of the COVID-19 pandemic.

Second, there is a dearth of evidence that shows how the coexistence of different clusters of mental disorders is related to COVID-19 clinical outcomes. Co-occurrence of different mental disorders are not infrequent events. Literature suggests that the co-occurrence of two or more conditions or disorders rates are very high in psychiatry and conform roughly to the rule of 50%. That is, about a half of people who meet the diagnostic criteria for one disorder meet the diagnostic criteria for a second disorder (28). Indeed, one 40-year longitudinal assessment among a cohort of 1,037 people in New Zealand reported that 85% of the participants with a mental disorder developed accumulated comorbid diagnoses by age 45 (25). It will be important for COVID-19 prevention and treatment to understand the impact of mental disorders (comorbidities) have on COVID-19 outcomes to inform effective surveillance and treatment.

Therefore, the current study aims to understand 1) how each cluster of pre-existing mental disorders (i.e., internalizing disorders, externalizing disorders, and thought disorders) is associated with COVID-19 clinical outcomes; and 2) how different clusters of mental disorders (i.e., internalizing & externalizing, internalizing & thought, externalizing & thought, internalizing & externalizing & thought) affect COVID-19 clinical outcomes. Using population-based EHR data from a statewide cohort of all confirmed and probable COVID-19 adult cases in South Carolina (SC), United States from March 6, 2020 to April 14, 2021, our study will provide database evidence to inform the healthcare delivery for people with pre-existing mental disorders during the COVID-19 pandemic.

## METHODS

### Data source

The data sources for the SC COVID-19 Cases (S3C) is described in a previous study conducted by our team (29). Mandatory reporting of COVID-19 cases to SC Department of Health and Environmental Control (DHEC) is required by SC Law and Regulations (30, 31). Specifically, all the key information of COVID-19 cases was collected through the SC statewide Case Report Form for SARS-CoV-2 infection (“Human Infection with 2019 Novel Coronavirus Case Report Form”, i.e., CRF) issued by SC DHEC. The key information of lab-confirmed and probable COVID-19 cases contained in the CRF includes the case classification and identification, hospitalization, ICU and death information, case demographics, clinical course, symptoms, past medical history, and social history. The criteria of case ascertainment follow the standardized surveillance case definition of COVID-19 (32). The EHR of COVID-19 patient were linked to all payer-based claims data through the SC Revenue and Fiscal Affairs (RFA) Office. SC state law mandates that the SC RFA receives reports on all diagnoses in International Classification of Disease (ICD) code format from all emergency departments, hospital inpatient facilities, ambulatory-care facilities, and outpatient surgery facilities in SC. The SC RFA office served as the honest data broker for data integration following the Health Insurance Portability and Accountability Act (HIPAA) regulations. De-identified data were then provided for our analysis. The study protocol was approved by the institutional review board in University of South Carolina and relevant SC state agencies. A total of 476,775 adult patients (age≥18 years old) with lab-confirmed and probable COVID-19 between March 06, 2020 and April 14, 2021 were included in the current study.

### Measures

#### Pre-existing mental disorders (independent variables)

Pre-existing mental disorder diagnoses from Jan 2, 2019 to Jan 14, 2021 were extracted from the patients’ healthcare utilization data via International Classification of Diseases [ICD] 10th edition [ICD-10] codes. The internalizing disorders included depression, generalized anxiety disorder, social phobia, simple phobia, agoraphobia, panic disorder, posttraumatic stress disorder, and eating disorders; the externalizing disorders included attention-deficit/hyperactivity disorder, conduct disorder, alcohol dependence, cannabis dependence, other drug dependence, and tobacco addition; and thought disorder were obsessive-compulsive disorder, mania, and schizophrenia (See Table 1 for ICD-10 code). A binary variable (Any clusters of mental disorders) was created based on the status of existing mental disorders. Patients who had at least one cluster of mental disorders before the COVID-19 diagnosis were defined as “Yes” for this variable and the rest were defined as “No”.

**Table 1.**
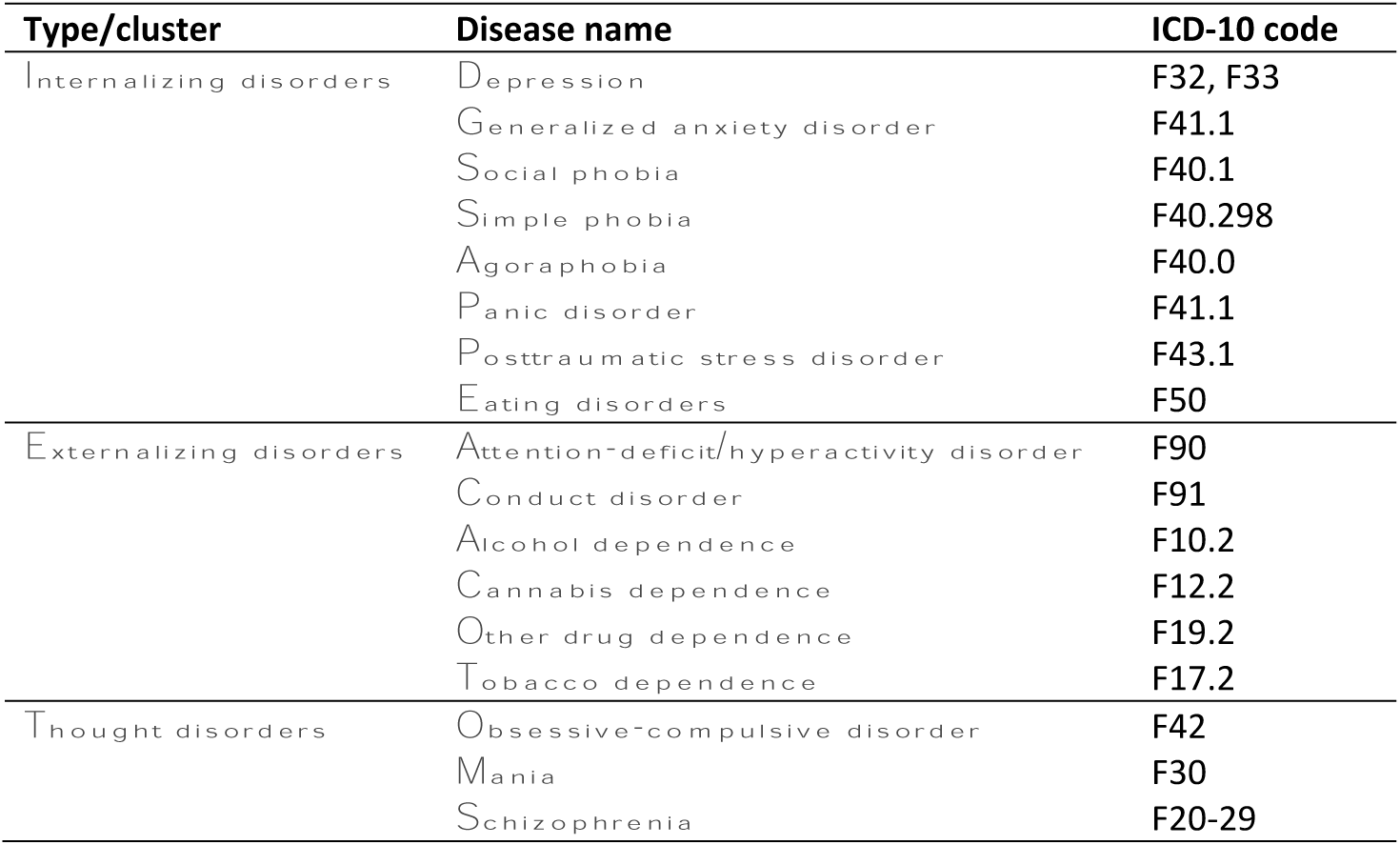
Summary of ICD-10 Codes used for Cluster Determination.

#### Clinical outcomes of COVID-19 (dependent variables)

Key COVID-19 clinical outcomes included severity, hospitalization, and death (29). COVID-19 severity was defined as asymptomatic, mild, and moderate/severe. COVID-19 patients who show no symptoms were categorized as asymptomatic; those who present any of various mild signs and symptoms of COVID-19 (e.g., fever, cough, sore throat, malaise, headache, muscle pain, nausea, vomiting, diarrhea, loss of taste and smell) were categorized as mild; and those with difficulty breathing or developed pneumonia or acute respiratory distress syndrome (ARDS) were categorized as moderate/severe. Hospitalization was assessed using response to the CRF question “Was the patient hospitalized?” (“yes”, “no”, or “unknown”). We then dichotomized the hospitalization (1=yes, 0=no or unknown). Death was measured by the question “Did the patient die as a result of this illness?” with the same three response items (“yes”, “no”, or “unknown”). It was dichotomized in the same way (1=death, 0=alive or unknown).

#### Covariates

Demographic characteristics included age at the time of COVID-19 diagnosis (18-49, ≥50), sex (female, male, and unknown), race (White, Black of African American, Asian, and other/unknown), and ethnicity (Hispanic or Latino, not Hispanic or Latino, and unknown). We created a binary variable of rural/urban residence according to the list of rural counties designated by the Office of Rural Health Policy (33). We also included smoking (never smoker, former smoker, current smoker, other/unknown), an established lifestyle factor associated with clinical outcomes of COVID-19 disease as one of covariates in statistical analysis (34).

### Statistical analysis

Descriptive statistics were computed for all study variables using means and standard deviations (SD) for the continuous variables and frequency counts and percentages for the categorical variables. Differences by each cluster of mental disorders by key COVID-19 outcomes (i.e., severity, hospitalization, and death) and covariates were tested using a t-test or Chi-square tests as appropriate. A Venn Diagram was used to illustrate patterns of co-occurrence of different clusters of mental disorders among the participants. We examined the association between pre-existing mental disorders cluster and the clinical outcomes of COVID-19 disease using logistic regression or multinomial logistic regressions for COVID-19 severity, hospitalization, and death, as appropriate separately. Models were fully adjusted for age, gender, race, ethnicity, residence, and smoking status. The models for COVID-19 hospitalization and death were also adjusted for the COVID-19 severity. To investigate if co-occurrence of different cluster of mental disorders associated with COVID-19 clinical outcomes, we further investigate the three-way interactions among three clusters of mental disorders in the final regression models. Forest plots were created based on the final regression models to virtually present the impacts to COVID-19 clinical outcomes by all the factors including pre-existing mental disorders, co-occurrence of different clusters of mental disorders, and other covariates. All the analyses were done with SAS, version 9.4. A p<0.05 was considered statistically significant for all 2-sided tests.

## RESULTS

### Characteristics of participants

Of the 476,775 adult COVID-19 patients included in the study, 421,475 patients had no recorded clinical diagnosis of a mental disorder and 55,300 patients who had at least one cluster of mental disorders before the COVID-19 diagnosis (Table 2). Among the overall sample, 58.3% were aged 18-49 years old (mean=45.7, SD=18.6), 52.9% were female, 47.6% were White, 20.4% were Black, 5.1% were Hispanic/Latino, and the majority were living in urban area (87.0%). Over half (54.0%) patients were identified as asymptomatic cases, 33.3% as mild cases, and 12.7% as moderate/severe cases. About 4.4% of all patients (n=20,995) were hospitalized and 1.9% (n=8,924) died due to COVID-19. Among the 55,300 patients who had at least one pre-existing cluster of mental disorders, 8.8% (n=4,848) were hospitalized and 3.9% (n=2,163) died due to COVID-19.

**Table 2.** Characteristics of the S3C population.

| Characteristics | Overall population | Any Cluster of Mental Disorders |  | Internalizing Disorders |  | Externalizing Disorders |  | Thought Disorders |  |
| --- | --- | --- | --- | --- | --- | --- | --- | --- | --- |
|  |  | No | Yes | No | Yes | No | Yes | No | Yes |
| **Age** |  |  |  |  |  |  |  |  |  |
| 18-49 | 277988<br>(58.31) | 249969<br>(59.31) | 28019<br>(50.67) | 263832<br>(59.46) | 14156<br>(42.78) | 259275<br>(58.1) | 18713<br>(61.27) | 276914<br>(58.4) | 1074<br>(41.09) |
| 50+ | 198787<br>(41.69) | 171506<br>(40.69) | 27281<br>(49.33) | 179849<br>(40.54) | 18938<br>(57.22) | 186960<br>(41.9) | 11827<br>(38.73) | 197247<br>(41.6) | 1540<br>(58.91) |
| **Gender** |  |  |  |  |  |  |  |  |  |
| Female | 252292<br>(52.92) | 219867<br>(52.17) | 32425<br>(58.63) | 229550<br>(51.74) | 22742<br>(68.72) | 237537<br>(53.23) | 14755<br>(48.31) | 251014<br>(52.94) | 1278<br>(48.89) |
| Male | 208956<br>(43.83) | 187738<br>(44.54) | 21218<br>(38.37) | 199609<br>(44.99) | 9347<br>(28.24) | 194059<br>(43.49) | 14897<br>(48.78) | 207712<br>(43.81) | 1244<br>(47.59) |
| Unknown/missing | 15527<br>(3.26) | 13870<br>(3.29) | 1657<br>(3) | 14522<br>(3.27) | 1005<br>(3.04) | 14639<br>(3.28) | 888<br>(2.91) | 15435<br>(3.26) | 92<br>(3.52) |
| **Race** |  |  |  |  |  |  |  |  |  |
| White | 226985<br>(47.61) | 200400<br>(47.55) | 26585<br>(48.07) | 208767<br>(47.05) | 18218<br>(55.05) | 213968<br>(47.95) | 13017<br>(42.62) | 225950<br>(47.65) | 1035<br>(39.59) |
| Black | 97066<br>(20.36) | 82522<br>(19.58) | 14544<br>(26.3) | 90289<br>(20.35) | 6777<br>(20.48) | 87672<br>(19.65) | 9394<br>(30.76) | 96192<br>(20.29) | 874<br>(33.44) |
| Other/Unknown | 149240<br>(31.3) | 135179<br>(32.07) | 14061<br>(25.43) | 141213<br>(31.83) | 8027<br>(24.26) | 141162<br>(31.63) | 8078<br>(26.45) | 148541<br>(31.33) | 699<br>(26.74) |
| **Ethnicity** |  |  |  |  |  |  |  |  |  |
| Not Hispanic or Latino | 285738<br>(59.93) | 250038<br>(59.32) | 35700<br>(64.56) | 263844<br>(59.47) | 21894<br>(66.16) | 266520<br>(59.73) | 19218<br>(62.93) | 284126<br>(59.92) | 1612<br>(61.67) |
| Hispanic or Latino | 24526<br>(5.14) | 23260<br>(5.52) | 1266<br>(2.29) | 23780<br>(5.36) | 746<br>(2.25) | 23839<br>(5.34) | 687<br>(2.25) | 24497<br>(5.17) | 29<br>(1.11) |
| Unknown/missing | 166511<br>(34.92) | 148177<br>(35.16) | 18334<br>(33.15) | 156057<br>(35.17) | 10454<br>(31.59) | 155876<br>(34.93) | 10635<br>(34.82) | 165538<br>(34.91) | 973<br>(37.22) |
| **Residence** |  |  |  |  |  |  |  |  |  |
|  |  | No | Yes | No | Yes | No | Yes | No | Yes |
| Rural | 61816<br>(12.97) | 53448<br>(12.68) | 8368<br>(15.13) | 57133<br>(12.88) | 4683<br>(14.15) | 56944<br>(12.76) | 4872<br>(15.95) | 61432<br>(12.96) | 384<br>(14.69) |
| Urban | 414959<br>(87.03) | 368027<br>(87.32) | 46932<br>(84.87) | 386548<br>(87.12) | 28411<br>(85.85) | 389291<br>(87.24) | 25668<br>(84.05) | 412729<br>(87.04) | 2230<br>(85.31) |
| **Smoking** |  |  |  |  |  |  |  |  |  |
| No | 71451<br>(14.99) | 61684<br>(14.64) | 9767<br>(17.66) | 64363<br>(14.51) | 7088<br>(21.42) | 67559<br>(15.14) | 3892<br>(12.74) | 70994<br>(14.97) | 457<br>(17.48) |
| Former smoker | 17274<br>(3.62) | 14392<br>(3.41) | 2882<br>(5.21) | 15445<br>(3.48) | 1829<br>(5.53) | 15724<br>(3.52) | 1550<br>(5.08) | 17207<br>(3.63) | 67<br>(2.56) |
| Current smoker | 8417 (1.77) | 5408<br>(1.28) | 3009<br>(5.44) | 7521<br>(1.7) | 896<br>(2.71) | 5580<br>(1.25) | 2837<br>(9.29) | 8325<br>(1.76) | 92<br>(3.52) |
| Other/Unknown | 379633<br>(79.63) | 339991<br>(80.67) | 39642<br>(71.69) | 356352<br>(80.32) | 23281<br>(70.35) | 357372<br>(80.09) | 22261<br>(72.89) | 377635<br>(79.64) | 1998<br>(76.43) |
| **COVID Severity/Syms** |  |  |  |  |  |  |  |  |  |
| No/asymptomatic | 257254<br>(53.96) | 226574<br>(53.76) | 30680<br>(55.48) | 239324<br>(53.94) | 17930<br>(54.18) | 239666<br>(53.71) | 17588<br>(57.59) | 255553<br>(53.9) | 1701<br>(65.07) |
| Mild | 158960<br>(33.34) | 143639<br>(34.08) | 15321<br>(27.71) | 150046<br>(33.82) | 8914<br>(26.94) | 150623<br>(33.75) | 8337<br>(27.3) | 158395<br>(33.41) | 565<br>(21.61) |
| Severe | 60561<br>(12.7) | 51262<br>(12.16) | 9299<br>(16.82) | 54311<br>(12.24) | 6250<br>(18.89) | 55946<br>(12.54) | 4615<br>(15.11) | 60213<br>(12.7) | 348<br>(13.31) |
| **COVID Hospitalization** |  |  |  |  |  |  |  |  |  |
| No | 455780<br>(95.6) | 405328<br>(96.17) | 50452<br>(91.23) | 426194<br>(96.06) | 29586<br>(89.4) | 427289<br>(95.75) | 28491<br>(93.29) | 453527<br>(95.65) | 2253<br>(86.19) |
| Yes | 20995 (4.4) | 16147<br>(3.83) | 4848<br>(8.77) | 17487<br>(3.94) | 3508<br>(10.6) | 18946<br>(4.25) | 2049<br>(6.71) | 20634<br>(4.35) | 361<br>(13.81) |
| **Death** |  |  |  |  |  |  |  |  |  |
| No | 467851<br>(98.13) | 414714<br>(98.4) | 53137<br>(96.09) | 436379<br>(98.35) | 31472<br>(95.1) | 438099<br>(98.18) | 29752<br>(97.42) | 465416<br>(98.16) | 2435<br>(93.15) |
| Yes | 8924 (1.87) | 6761<br>(1.6) | 2163<br>(3.91) | 7302<br>(1.65) | 1622<br>(4.9) | 8136<br>(1.82) | 788<br>(2.58) | 8745<br>(1.84) | 179<br>(6.85) |
|  |  | No | Yes | No | Yes | No | Yes | No | Yes |

**Note:** The results for Asian group were not reported due to policy. Chi-square tests for all the characteristics variables are all significant (i.e., $P < .05$ ).

### Pre-existing mental disorders and clinical outcomes of COVID-19

Of the 55,300 patients with pre-existing mental disorders, 23,410 (42.3%) had internalizing disorders only, 21,200 (38.3%) had externalizing disorders only, and 618 (1.1%) had thought disorders only. The number of individuals with a single cluster or multiple clusters of mental disorders is presented in a Venn diagram (Figure 1). There were 8,076 (14.6%) patients who had both internalizing and externalizing disorders, 732 (13.2%) had both internalizing and thought disorders, and 388 (0.7%) had externalizing and thought disorders. The number of patients having all three clusters of internalizing, externalizing, and thought disorders was 876 (1.6%).

**Figure 1.**
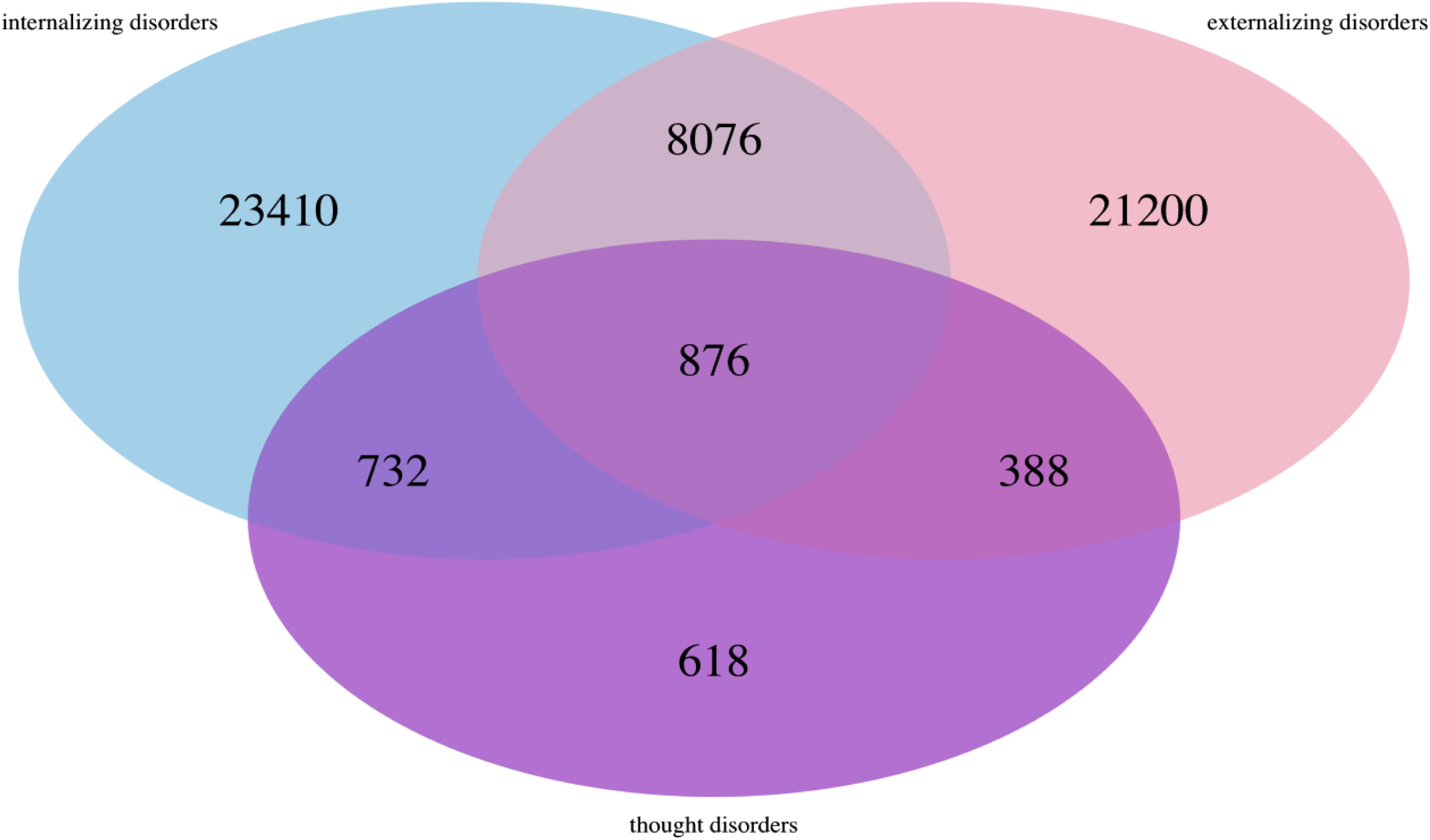
Venn Diagram for comorbidity of mental disorders among the S3C population: Sample in each subgroup. **Note:** Not any mental disorders=421,475; internalizing disorders only =23,410; externalizing disorders only =21,200; thought disorders only=618; internalizing and externalizing disorders=8,076; internalizing and thought disorders=732; externalizing and thought disorders=388; internalizing, externalizing, and thought disorders=876.

Generally, we observed significant differences in age, gender, race, ethnicity, residence, and smoking status between participants with and without pre-existing mental disorders (Table 2). For example, there was a higher proportion of non-Hispanic/Latino among participants with pre-existing mental disorders, compared with those without these conditions. However, the patterns of the socio-demographic difference between participants with and without pre-existing mental disorders varied by the cluster of mental disorders. Higher proportions of participants with internalizing disorders were older (≥50 years), female, White, non-smokers, and lived in rural areas. Higher proportions of participants with externalizing disorders were males, Black/African American, former/ current smokers, and lived in rural areas. We found higher proportions of participants with thought disorders were older (≥50 years), Black/African American, non-smokers, and lived in rural areas.

Regarding the clinical outcomes of COVID-19, there was a higher proportion of hospitalization for COVID-19 among participants with pre-existing mental disorders, compared to those without respective cluster of mental disorders (Internalizing disorders: 10.6% vs. 3.9%; Externalizing disorders: 6.7% vs. 4.3%; Thought disorders: 13.8% vs. 4.4%; Any cluster of mental disorders: 8.8% vs. 3.8%) and COVID-19-related deaths (Internalizing disorders: 4.9% vs. 1.7%; Externalizing disorders: 2.6% vs. 1.8%; Thought disorders: 6.9% vs. 1.8%; Any cluster of mental disorders: 3.9% vs. 1.6%). The association between pre-existing mental disorders and COVID-19 severity was more complicated. In general, participants with pre-existing mental disorders had higher proportions of being identified as asymptomatic (Internalizing disorders: 54.2% vs. 53.9%; Externalizing disorders: 57.6% vs. 53.7%; Thought disorders: 65.1% vs. 53.9%; Any cluster of mental disorders: 55.5% vs. 53.8%) and severe cases (Internalizing disorders: 18.9% vs. 12.2%; Externalizing disorders: 15.1% vs. 12.5%; Thought disorders: 13.3% vs. 12.7%; Any cluster of mental disorders: 16.8% vs. 12.2%). However, they showed a lower proportion to have mild status (Internalizing disorders: 26.9% vs. 33.8%; Externalizing disorders: 27.3% vs. 33.8%; Thought disorders: 21.6% vs. 33.4%; Any cluster of mental disorders: 27.7% vs. 34.1%).

### Regression analysis results

The multivariate results suggest an elevated risk of COVID-19-related hospitalization and death among participants with pre-existing mental disorders adjusting for key socio-demographic covariates (i.e., age, gender, race, ethnicity, residence, smoking) (Table 3). For COVID-19 hospitalization, the adjusted OR of having internalizing disorders, externalizing disorders, and thought disorders was 2.27 (95%CI 2.157, 2.386), 1.49 (95%CI 1.393,1.599), and 4.41 (95%CI 3.516, 5.534), respectively. Similarly, for COVID-19-related death, the adjusted OR of having internalizing disorders, externalizing disorders, and thought disorders was 2.25 (95%CI 2.109, 2.407), 1.33 (95%CI 1.199,1.472), and 4.48 (95%CI 3.435,5.839), respectively. However, pre-existing mental disorders was associated with decreased risk of mild and severe cases. Specifically, having internalizing disorders (OR=0.65, 95%CI 0.624,0.671), externalizing disorders (OR=0.64, 95%CI 0.611,0.660), and thought disorders (OR=0.54, 95%CI 0.434,0.680) were negatively associated with mild cases. Having externalizing disorders (OR=0.78, 95%CI 0.742,0.818) and thought disorders (OR=0.75, 95%CI 0.565, 0.990) was negatively associated with severe cases, but having internalizing disorders (OR=1.09, 95%CI 1.042,1.135) was positively associated with severe cases.

**Table 3.** Regression model results for COVID-19 outcomes adjusting for covariates in the S3C population.

| Items | Severity (mild vs no) | Severity (severe vs no) | Hospitalization | Death |
| --- | --- | --- | --- | --- |
|  | OR (95%CI) | OR (95%CI) | OR (95%CI) | OR (95%CI) |
| age 50+ vs 18-49 | **0.640 (0.630,0.650)** | **0.711 (0.696,0.726)** | **8.280 (7.952,8.622)** | **30.488 (27.395,33.931)** |
| Gender Male vs Female | **0.912 (0.898,0.926)** | **0.827 (0.810,0.844)** | **1.505 (1.459,1.553)** | **1.579 (1.510,1.651)** |
| Gender unknown/missing vs Female | **0.920 (0.875,0.967)** | **0.844 (0.786,0.907)** | **0.754 (0.665,0.854)** | **0.724 (0.604,0.867)** |
| Race Asian vs White | 1.059 (0.980,1.144) | 0.981 (0.879,1.095) | **1.357 (1.124,1.639)** | 1.067 (0.791,1.440) |
| Race Black vs White | **0.788 (0.774,0.803)** | **0.865 (0.844,0.886)** | **2.049 (1.979,2.121)** | **1.335 (1.268,1.405)** |
| Race Other/Unknown vs White | **0.253 (0.247,0.259)** | **0.264 (0.255,0.274)** | **0.860 (0.811,0.912)** | **0.813 (0.752,0.880)** |
| Ethnicity Hispanic or Latino vs Not Hispanic or Latino | **2.065 (1.995,2.137)** | **2.440 (2.332,2.553)** | **1.454 (1.345,1.571)** | **0.858 (0.745,0.988)** |
| Ethnicity Unknown/missing vs Not Hispanic or Latino | **0.291 (0.285,0.297)** | **0.276 (0.267,0.285)** | **0.679 (0.645,0.714)** | **0.876 (0.819,0.937)** |
| Urban vs Rural | **1.063 (1.039,1.086)** | **1.098 (1.066,1.131)** | **0.876 (0.841,0.913)** | **0.808 (0.764,0.856)** |
| Smk Current smoker vs No | 0.970 (0.898,1.046) | 1.021 (0.941,1.108) | **0.494 (0.442,0.552)** | **0.490 (0.408,0.589)** |
| Smk Former smoker vs No | **1.494 (1.400,1.595)** | **1.996 (1.866,2.136)** | **0.888 (0.839,0.941)** | **0.698 (0.638,0.763)** |
|  | OR (95%CI) | OR (95%CI) | OR (95%CI) | OR (95%CI) |
| Smk<br>Other/Unknown vs<br>No | **0.103 (0.101,0.106)** | **0.066 (0.064,0.068)** | **0.795 (0.766,0.825)** | **0.866 (0.820,0.915)** |
| internalizing_disord<br>Yes vs No | **0.647 (0.624,0.671)** | **1.088 (1.042,1.135)** | **2.269 (2.157,2.386)** | **2.253 (2.109,2.407)** |
| externalizing_disord<br>Yes vs No | **0.635 (0.611,0.660)** | **0.779 (0.742,0.818)** | **1.493 (1.393,1.599)** | **1.329 (1.199,1.472)** |
| thought_disorders<br>Yes vs No | **0.543 (0.434,0.680)** | **0.748 (0.565,0.990)** | **4.411 (3.516,5.534)** | **4.479 (3.435,5.839)** |
| Internal & external<br>v.s. No | **0.439 (0.412,0.467)** | **0.887 (0.826,0.952)** | **2.496 (2.284,2.728)** | **2.022 (1.782,2.295)** |
| Internal & thoughts<br>vs. No | **0.510 (0.415,0.626)** | 0.961 (0.760,1.214) | **3.339 (2.637,4.228)** | **3.166 (2.364,4.240)** |
| External & thoughts<br>vs. No | **0.447 (0.336,0.595)** | **0.493 (0.333,0.730)** | **3.291 (2.288,4.733)** | **3.671 (2.398,5.621)** |
| Internal & external<br>& thought vs. No | **0.294 (0.240,0.359)** | **0.489 (0.384,0.622)** | **3.350 (2.604,4.310)** | **1.657 (1.093,2.512)** |
| sympt Mild vs<br>asympt | n/a | n/a | **1.256 (1.199,1.315)** | **0.756 (0.707,0.809)** |
| sympt Severe vs<br>asympt | n/a | n/a | **10.007 (9.587,10.445)** | **4.751 (4.479,5.040)** |

Note: Bold OR indicate P-value <.05.

Co-occurrence of any two clusters was positively associated with COVID-19 hospitalization and COVID-19-related death (See the forest plot in Figure 2 and Table 3). Specifically, compared to those with no pre-existing mental disorders, the odds of being hospitalized was 1.50 times greater among patients with internalizing and externalizing disorders (OR=2.50, 95%CI 2.284, 2.728), 2.34 times greater among patients with internalizing and thought disorders (OR=3.34, 95%CI 2.637, 4.228), 2.29 times greater among patients with externalizing and thought disorders (OR=3.29, 95%CI 2.288, 4.733), and 2.35 times greater among patients with three clusters of mental disorders (OR=3.35, 95%CI 2.604, 4.310). Similarly, compared with the COVID patients with no pre-existing mental disorders, the odds of having COVID-19-related death was 1.02 greater among patients with internalizing and externalizing disorders (OR=2.02, 95%CI 1.782, 2.295), 2.17 greater among patients with internalizing and thought disorders (OR=3.17, 95%CI 2.364, 4.240), 2.67 greater among patients with externalizing and thought disorders (OR=3.67, 95%CI 2.398, 5.621), and 67% greater among patients with a co-occurrence of three clusters of mental disorders (i.e., internalizing, externalizing and thought disorders; OR=1.66, 95%CI 1.093, 2.512). Co-occurrence of any two clusters was negatively related with mild cases. The odds of having mild symptoms decreased 56% among patients with internalizing and externalizing disorders (OR=0.44, 95% CI 0.412, 0.467), 49% for patients with internalizing and thoughts disorders (OR=0.51, 95% CI 0.415, 0.626), 55% for patients with externalizing and thoughts disorders (OR=0.45, 95% CI 0.336, 0.595), and 71% for patients with three clusters of mental orders (OR=0.29, 95% CI 0.240, 0.359). In addition, the odds of having severe symptoms decreased 11% for patients with internalizing and externalizing disorders (OR=0.89, 95%CI 0.826, 0.952), 51% among patients with externalizing and thought disorders (OR=0.49, 95%CI 0.333, 0.730) and 51% for patients with three clusters of mental disorders (OR=0.49, 95%CI 0.384, 0.622).

**Figure 2.**
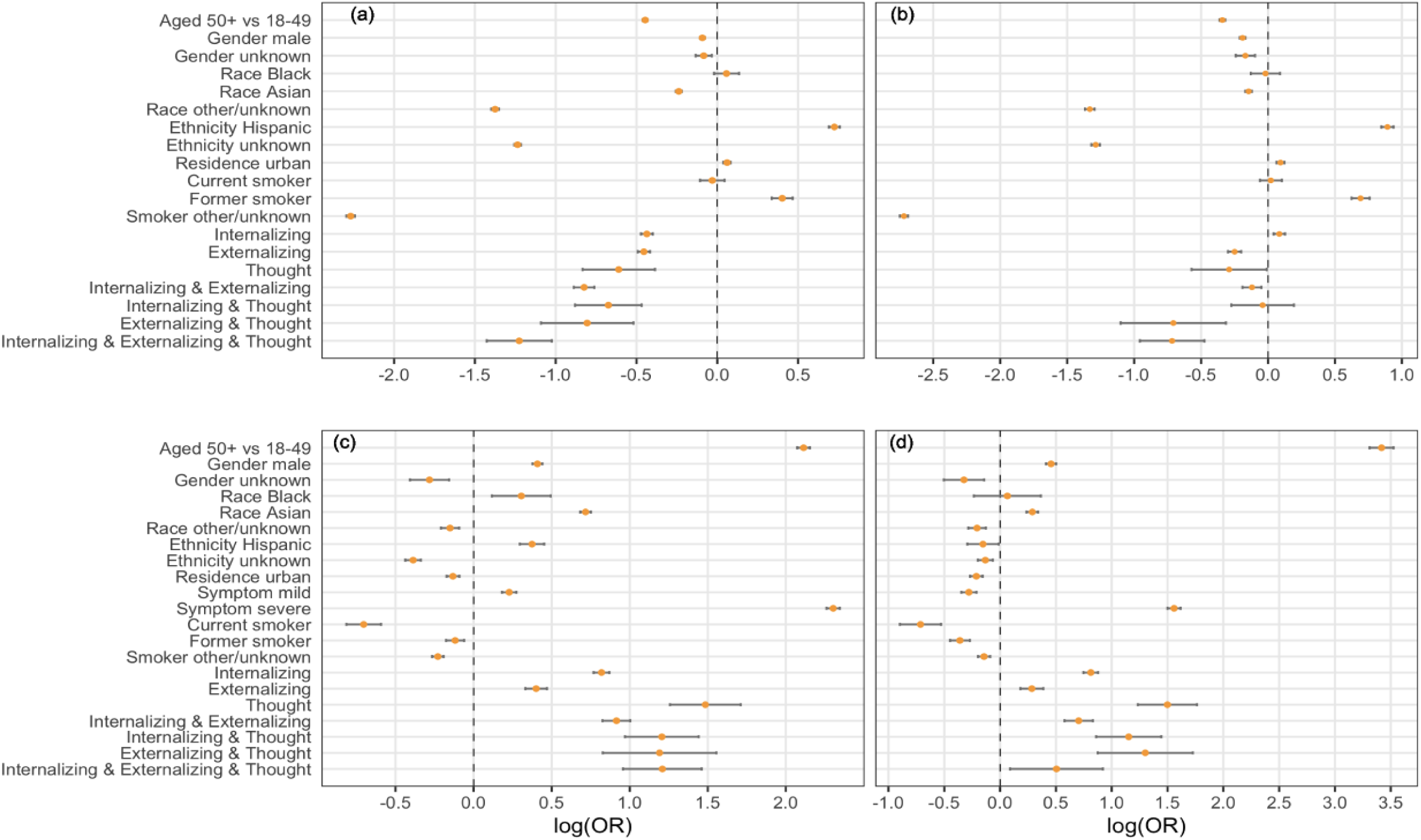
Forest plot of final regression models. **Note:** Logarithm of odds ratio was used in developing the forest plots given large value of some odds ratios. We then use zero instead of one as the criteria of significance. (a) severity (mild vs asymp); (b) severity (severe vs asymp); (c) hospitalization (yes vs no); (d) death (yes vs no).

## DISCUSSION

Using a psychopathology structure model of mental disorders (i.e., the three-factor model) and leveraging the EHR data for state-level cohort of COVID-19 patients in SC, United States, we have explored the association between pre-existing mental disorders and severity of COVID-19 from a new perspective. That is, we examined how each cluster of mental disorders (i.e., internalizing disorders, externalizing disorders, and thought disorders) could affect COVID-19 clinical outcomes (i.e., severity, hospitalization, and death) and how the co-occurrence of multiple clusters of mental disorders associates with these clinical outcomes after controlling for other key sociodemographic confounders (e.g., age, race, ethnicity, urban/rural residence).

Our findings are aligned with existing evidence of a positive association between pre-existing mental disorders and COVID-19 related hospitalization and death. Several studies have posited potential reasons and interpretations regarding the underlying mechanism of pre-existing mental disorders as a risk factor of COVID-19 clinical outcomes, including immune dysregulation processes, genetic predisposition towards psychiatric disorders, and health-compromising lifestyle (20). The activation of the endocrine stress axis at different levels, such as hypothalamic-pituitary-adrenal (HPA) axis which can lead to altered glucocorticoids and suppressed cell-mediated and humoral immunity is widely reported among people with mental disorders (35-38). The immune dysfunction, subsequently, contributes to high risk of SARS-CoV-2 infection. The inflammatory cytokine overproduction induced by glucocorticoid receptor resistance may be responsible for the damage of the lungs and correlated with disease deterioration and fatal COVID-19 (39-41). Recent studies also demonstrate a strong genetic link between psychiatric disorders and general infections including COVID-19. Health behavioral changes in response to stress (e.g., altered diet and exercise patterns, smoking and alcohol abuse) may also explain the observed associations between mental disorders and COVID-19 clinical outcomes (42).

In addition, we discovered that pre-existing thought disorders (e.g., mania, schizophrenia, obsessive-compulsive disorders [OCD]) may be related to COVID-19 hospitalization and death in the greatest magnitude. Thought disorders are usually considered as severe mental disorders. For example, mania occurs in the context of bipolar disorder and schizophrenia. A recent systematic review and meta-analysis on the association between mental disorders and COVID-19 death reports that people with severe mental disorders (defined as schizophrenia and/or bipolar disorders) have the highest risk of COVID-19 death (OR: 1.67, 95% CI, 1.02-2.73) (20). One national-level study conducted in South Korea suggests that severe mental disorders (defined as non-affective or affective disorders with psychotic features) are more likely to be associated with severe clinical outcomes (admission to the ICU, invasive ventilation, death) compared to other mental disorders (OR: 2.27, 95% CI, 1.50-3.41) (16). Our findings are consistent with extant literature suggesting that patients with severe mental disorders such as thought disorders face higher risk of worse COVID-19 clinical outcomes. The explanation from immunological perspective includes variation of human leukocyte antigen (which predominately regulates viral infection and may contribute to differences in the immune response to COVID-19) and abnormal cytokine levels among patients with schizophrenia and bipolar disorders (43-48). In addition, people with thought disorders are likely to need care and support of caregivers to follow healthcare and COVID-19 mitigation measures such as wearing face masks and social distancing. Some prevention recommendations, such as hand washing, can reinforce the irrational beliefs of patients with OCD (48). The limited availability of caregivers and tailored COVID-19 mitigation strategies during the pandemic makes this group more vulnerable to COVID-19.

We found that people with co-occurrence of any two clusters of mental disorders have elevated risk of COVID-19-related hospitalization and death compared to those with a single cluster (either internalizing or externalizing disorder). Given that having pre-existing thought disorders is a high-risk factor of worse COVID-19 clinical outcomes, it is reasonable that co-occurrence of thought disorders and either internalizing disorders or externalizing disorders indicates more vulnerability compared to the single cluster of pre-existing mental disorders. Notably, the co-occurrence of internalizing and externalizing disorders has increased risk of hospitalization and death caused by COVID-19 compared to a single condition. Internalizing disorders, like depression and anxiety disorders have been found to be associated with immune-inflammatory disturbances, which may contribute to severe clinical outcomes of COVID-19(49, 50). The lifestyle changes following the occurrence of internalizing disorders, such as smoking and alcohol abuse, may also contribute to altered risk to COVID-19 infection and worse outcomes. Externalizing disorders, especially long-term use of tobacco, alcohol, and other drugs, are also closely related to cardiovascular, pulmonary, and metabolic diseases, which are high-risk preconditions for COVID-19 outcomes(51, 52). In addition, some studies indicate a genetic association between psychiatric disorder and COVID-19(53). For example, one genome-wide association study suggests that genetic liability to depression and substance misuse is associated with severe COVID-19 outcomes (hospitalization or death)(54).

Although our results suggest that pre-existing mental disorders are risk factors of COVID-19-related hospitalization and death, it is interesting that pre-existing mental disorders may be associated with decreased risk of mild cases. Extant literature suggests that pre-existing chronic illness and therapeutic options may affect the prognosis of COVID-19 and have a possible protective effect against COVID-19. For example, allergic sensitization in asthma is related to lower expression of angiotensin-converting enzyme (ACE)2 receptors showing a potential protective effect(55). In addition, the use of inhaled corticosteroids is generally safe and associated with decreased risk of hospitalization(56). However, there is a dearth of evidence regarding whether treatment for mental disorders affect prognosis of COVID-19. Given the high rate of comorbidity between mental disorders and chronic somatic disease, the pre-existing chronic conditions and the treatment among these patients could be complicated. Further studies are needed to identify potential confounders and explain the “protective effect”(56). In addition, the high-risk drug-drug interactions (DDIs) that may occur in COVID-19 treatment accompanied by psychotropic drug prescription warrants multidisciplinary study engaging both psychiatrists and infectious disease physicians(57).

The strengths of the current study include categorizing mental disorders guided by a psychopathology structure model, a statewide cohort, being representative of both impatient and outpatient COIVD-19 cases and generating real-world evidence. Utilization of the statewide standardized case report form enables us to systematically collect comprehensive information on COVID-19 clinical outcomes. Our findings need to be interpreted with caution due to the following limitations: First, our mental health data comes from a health utilization dataset. Therefore, we were not able to retrieve information of people who had mental disorders but failed to access to healthcare system due to any individual or structural level barriers. There is also missing information in the COVID-19 data. The missing data may impede the robustness of our findings. Second, the study is subject to common limitations of using ICD10 code to define mental health conditions. As other EHR based studies suggest, the quality of raw data may influence the validity of our results. Finally, we did not differentiate the severity category of the mental disorders within the same cluster. For example, we did not explore if patients with “acute” disorders have any different risk of worse COVID-19 outcomes compared to those with “recurrent” (more severe mental illness) disorders.

Despite these limitations, the current study sheds lights on impacts of different clusters of mental disorders on COVID-19 clinical outcomes and the risk of presenting severe COVID-19 clinical outcomes among patients with co-occurrence of multiple clusters of mental disorders using a large statewide and real-world dataset. Our findings have significant implications for improving surveillance and triage in COVID-19 treatment considering that a high prevalence of psychiatric disorders (20.6% as of 2019) exists in the general population (58). First, it is critical to identify vulnerable subgroups in COVID-19 treatment. Pre-existing thought disorders and co-occurrence of multiple clusters of mental disorders seem to be independent risk factors of COVID-19-related hospitalization and death controlling other background characteristics, which will assist triage and medical resource allocation so that the healthcare providers can pay attention to the most vulnerable subgroup and prevent the disease deterioration among them. Specific strategies may include providing triage at admission, reducing mental health stigma in clinical settings, and improving access to health resources in time to prevent patients with mental disorders from late admission with serious conditions.

Second, sociodemographic profiles of patients with comorbidity of both COVID-19 and mental disorders warrants further exploration. Our findings show that higher proportions of patients with thought disorders were older (≥50 years), Black/African American, and lived in rural area. Mounting evidence indicates mental health disparities in the United States with racial/ethnic minorities and rural people being disadvantaged in medical and mental health settings. Rural areas persistently face a lack of mental health professionals and mental health service infrastructure (59). Compared to Whites, African Americans have less access to mental health services and less likely to utilize necessary mental healthcare. They are less satisfied with professional mental health services and have higher rates of dropout from these services (60). Furthermore, this group faces higher risk of COVID-19 infection, endures more barriers to access to health resources, and shows disproportionally high morbidity and mortality of COVID-19. More empirical evidence and policy studies are needed to illustrate and address the intersection between mental health, COVID-19, and social deprivation/vulnerability from the perspective of health equity and structural racism.

## CONCLUSION

In summary, the current study suggests that people with mental disorders, especially thought disorders and co-occurrence of multiple clusters should been identified as high-risk population for severe consequences of COVID-19, requiring enhanced preventive, triage, and treatment strategies. Future studies need to further differentiate the impacts of mental disorders on COVID-19 clinical outcomes by severity/stage of the illness (e.g., acute vs. stabilized); include a comprehensive set of social determinates of health in the analysis; and explore the interaction between different clusters of mental disorders and pre-existing somatic conditions.

## Data Availability

All data produced in the present study are available upon reasonable request to the authors

## Notes

### Competing Interest Statement

The authors have declared no competing interest.

### Funding Statement

This study was funded by NIH R01AI127203-4S1

### Author Declarations

The study protocol was approved by the institutional review board in University of South Carolina and relevant SC state agencies.

